# Factors associated with COVID-19 vaccine receipt at two integrated healthcare systems in New York City: A Cross sectional study of healthcare workers

**DOI:** 10.1101/2021.03.24.21253489

**Authors:** Kristin Oliver, Anant Raut, Stanley Pierre, Leopolda Silvera, Alexander Boulos, Alyssa Gale, Aaron Baum, Ashley Chory, Nichola Davis, David D’Souza, Amy Freeman, Crispin Goytia, Andrea Hamilton, Carol Horowitz, Nadia Islam, Jessica Jeavons, Janine Knudsen, Sheng Li, Jenna Lupi, Roxanne Martin, Sheela Maru, Ismail Nabeel, Dina Pimenova, Anya Romanoff, Nina Schwalbe, Nita Vangeepuram, Rachel Vreeman, Joseph Masci, Duncan Maru

**Author notes:** Corresponding author Kristin Oliver.

## Abstract

**Objectives:** To examine factors associated with COVID-19 vaccine receipt among healthcare workers, including healthcare worker job type, race, and gender, as well as the role of vaccine confidence in decisions to vaccinate, and to better understand specific concerns related to COVID-19 vaccination among healthcare workers.

**Design:** Cross-sectional anonymous survey among front-line, support service, and administrative healthcare workers.

**Setting:** Two large integrated healthcare systems (one private and one public) in New York City during the initial rollout of the COVID-19 vaccine among healthcare workers.

**Participants:** 1,933 healthcare workers, including nurses, physicians, allied health professionals, environmental services staff, researchers, and administrative staff.

**Main Outcome Measures:** The primary outcome was COVID-19 vaccine receipt during the initial rollout of the vaccine among healthcare workers.

**Results:** Among 1,933 healthcare workers who had been offered the vaccine, 81% had received the vaccine at the time of the survey. Receipt was lower among Black (58%) compared with White (91%) healthcare workers; and lower among Hispanic (69%) compared with non-Hispanic (84%) healthcare workers. Among healthcare workers with concerns about COVID-19 vaccine safety, 65% received the vaccine. Among healthcare workers who agreed with the statement that the vaccine is important to protect family members, 86% were vaccinated. Of those who disagreed, 25% received the vaccine. Across all participants, 27% expressed concern about being experimented on with the COVID-19 vaccine. In a multivariable analysis, concern about being experimented on with the COVID-19 vaccine, concerns about COVID-19 vaccine safety, lack of influenza vaccine receipt, disagreeing that COVID-19 vaccination is important to protect family members, and Black race were independently associated with COVID-19 vaccine non-receipt. Over 70% of all healthcare workers responded that they had been approached for vaccine advice multiple times by family, community members, and patients.

**Conclusions:** Our data demonstrated high overall receipt among healthcare workers. Even among healthcare workers with concerns about COVID-19 vaccine safety, side effects, or being experimented on, over 50% received the vaccine. Attitudes around the importance of COVID-19 vaccination to protect others played a large role in healthcare workers’ decisions to vaccinate. We observed striking inequities in COVID-19 vaccine receipt, particularly affecting Black and Hispanic workers. Further research is urgently needed in developing strategies with healthcare workers to address issues related to vaccine equity and uptake in the context of systemic racism and barriers to care. This is particularly important given the influence healthcare workers have in vaccine decision-making conversations in their communities.

**SUMMARY BOXES:** 

**What is already known?:**

- High uptake of effective COVID-19 vaccines among healthcare workers is critical to pandemic response.
- In studies of potential COVID-19 vaccine acceptance prior to COVID-19 vaccine availability, people who identified as Black were less likely to indicate they would accept the vaccine.
- Understanding reasons why some healthcare workers chose not to get the COVID-19 vaccine will help us develop interventions to improve COVID-19 vaccine confidence among healthcare workers and in their communities.

**What this study adds:**

- We demonstrate high receipt of COVID-19 vaccines in the initial rollout among healthcare workers.
- Attitudes around the importance of COVID-19 vaccination to protect others played a large role in healthcare workers’ decisions to vaccinate.
- We observed substantially lower rates of receipt among Black and Hispanic healthcare workers, independent of differences in vaccine-related beliefs. A quarter of healthcare workers expressed concerns about being experimented on. These results suggest systemic racism may be a critical barrier to equitable vaccination.
- Our results highlight that healthcare workers of all types, including those with non-patient-facing roles, play an important role as sources of COVID-19 vaccine information in their communities.

## Introduction

The equitable and widespread uptake of effective vaccines against SARS-CoV-2 will be critical to control the COVID-19 pandemic. Approval of the first COVID-19 vaccines in the United States was preceded by months of intense public and political discourse on potential vaccine efficacy and safety. Throughout 2020, estimates of COVID-19 vaccination intent in the general adult population ranged from less than half to approximately three-quarters, and pointed to an urgent need to address vaccine confidence.^1^ Members of groups affected by systemic racism suffered disproportionate morbidity and mortality during the COVID-19 pandemic.^2-7^ These groups, in particular Blacks, have long suffered gross abuses and injustices in healthcare. Partly as a result, members of marginalized groups have expressed lower intent to receive COVID-19 vaccines.^2,8,9^

In December 2020, the first COVID-19 vaccines were authorized for emergency use in the United States and healthcare workers were among the first groups to be offered the vaccines.^10^ Recent working papers have highlighted the need to effectively engage local communities in COVID-19 vaccination^11^ and to equitably distribute the vaccines.^12^

Globally, local healthcare workers are some of the most trusted and influential professionals in individual and family decisions around vaccination.^13,14^ Beyond their work roles, healthcare workers are also influential members of the communities in which they live. Vaccine hesitancy is prevalent among healthcare workers globally.^15^ The World Health Organization (WHO) emphasizes that targeted discussions and engagement with healthcare providers will be essential to obtaining widespread vaccination confidence, as they will be the first ones expected to get the vaccine, and they will be on the frontlines facing questions from the public.^16^

Our group at the COVID-19 Unit for Research at Elmhurst (CURE-19) partnership between NYC Health & Hospital Corporation’s Elmhurst and Queens Hospital Centers and the Arnhold Institute for Global Health at Mount Sinai, recognized that the COVID-19 pandemic and vaccine confidence are going to be long-standing public health issues over the coming decade. The healthcare systems that our partnership represent cover over 7 million patient visits annually and 80,000 healthcare workers in neighborhoods with some of the highest linguistic, cultural, and ethnic diversity in the United States. Here, we aimed to examine the demographic, work role, and vaccine-related belief factors associated with COVID-19 vaccine receipt among healthcare workers during the initial rollout of the COVID-19 vaccine in this highly diverse region that has been heavily impacted by the pandemic.

## Methods

### Setting

We conducted an online cross-sectional survey of healthcare workers at two large integrated healthcare systems (one public and one private) in New York City. COVID-19 vaccines were available in New York City beginning December 14, 2020. Based on New York State guidelines, healthcare workers at high risk for exposure were in the first COVID-19 vaccine eligibility category. Vaccine eligibility expanded throughout our data collection time period.

For participant eligibility, we defined healthcare worker broadly to include physicians, nurses, allied health professionals, advanced practice providers, environmental services workers, community-based providers, and researchers/educators. We recruited participants through use of hospital listservs, newsletters, and emails, and through distributing posters and flyers onsite at locations across the health systems. The survey was available in English, Spanish, Bangla, Mandarin, Nepali, and Haitian Creole. An incentive for a chance to win one of ten $50 cash prizes was offered to the survey participants. The survey was available electronically via REDCap and by paper upon request.

The online form included exclusively forced choice questions, except for two open ended questions with free text answers included at the end of the survey. As such there were no missing data. We also piloted paper surveys, which were completed by nine participants. We did not include them in the primary analysis, as the data were incomplete or clearly inaccurate.

Data collection ran from December 23, 2020, to February 16, 2021, corresponding to the first two months of COVID-19 vaccine rollout.

### Main Measures

We asked respondents if they had been offered a COVID-19 vaccine by the time of the survey. For those who had been offered a vaccine, we asked whether they had received the vaccine, not received the vaccine, or “planned to get it but had not scheduled or gone for the vaccine yet”. For participants who had not been offered the vaccine, we asked them if when a vaccine was offered to them, they intended to “get one as soon as possible”, “delay getting it for a few months”, or “never get one”.

To measure general vaccine confidence, we used the Vaccine Confidence Index (VCI), which was developed and validated by The Vaccine Confidence Project to measure “individual perceptions on the safety, importance, effectiveness, and religious compatibility of vaccines”.^17-19^ The three statements are: “Overall, I think vaccines are safe”, “I think vaccines are important for children to have”, and “Overall, I think vaccines are effective”. For computing the Vaccine Confidence Index as a numerical score, we assigned numerical values to responses to each of the three questions (strongly agree = 5, somewhat agree = 4, neither agree nor disagree = 3, somewhat disagree = 2, strongly disagree = 1). While there is, as yet, no standardized way of categorizing responses, we used existing literature^20^ and visual inspection of the data to assign VCI scores less than 9 as low vaccine confidence.

We further adapted the VCI questions to explore COVID-19 specific disease risk appraisal, vaccine benefits, and vaccine harms (see Supplementary Materials). We also asked about influenza vaccine receipt in the current 2020-2021 influenza season, and about social media exposures.

We included questions on trusted sources for COVID-19 vaccine advice, information seeking behavior, and how often coworkers, friends and family members asked for respondents’ opinions on COVID-19 vaccination. To better understand specific concerns related to COVID-19 vaccination, we included two optional open-ended questions at the end of the survey inviting participants to share their thoughts and suggestions and inquiring about what tools and resources would help them provide information about COVID-19 with more confidence. We collected demographic and occupational variables including race, ethnicity, age, gender, job role and involvement in direct patient care

### Quantitative Analytical Methods

We report COVID-19 vaccine receipt by demographic and occupational factors, general vaccine confidence, influenza vaccine receipt and COVID-19 vaccine attitudes. We expressed results in the form of means, proportions, and 95% confidence intervals. We conducted bivariate analyses using Chi-squared tests for categorical variables and Wilcoxon rank-sum tests for the continuous variable Vaccine Confidence Index, with significance set at α=0.05. For this analysis focused specifically on vaccine receipt (rather than intent) as the primary outcome, respondents who had not yet received the vaccine but planned to get it were included in the “no” category.

We developed a multivariable logistic regression model to assess the adjusted and relative contributions of demographic variables and vaccine confidence on COVID-19 vaccine receipt when offered. For the logistic regression model, we utilized the Lasso procedure,^21^ in R Studio, Version 1.4.1103, which uses regularization, cross-validation and penalization to identify important predictor variables and improve the interpretability and predictive accuracy of the final statistical model. We used 50% of the dataset for training and 50% for testing. Independent variables inputted into the model included demographic variables, occupational variables, and workers’ perceptions of COVID-19 vaccine benefits, harms, and disease risk. In a supplementary table, we report which variables were retained (had positive coefficient weights), not retained (had zero weight), and their coefficients (transformed into an odds ratio) in the final adjusted model. For generating odds ratios and confidence intervals in the final model, we utilized the glm procedure in R.

### Qualitative Analytical Methods

We analyzed the free-text survey response data through summary descriptive statistics. Review of survey response transcripts allowed for preliminary thematic analysis to formulate detailed narratives. Open coding of survey transcripts was completed by two team members (AC, LS), which led to an initial set of codes, which were refined by a third member (RM). These were organized according to themes that emerged from the data and by the research premise and conceptual model. We generated a codebook of code definitions and examples. We completed coding of all transcripts completed by two team members, using Dedoose software (SocioCultural Research Consultants, LLC, Manhattan Beach, California, USA). We formulated the final set of overarching themes in discussion with the full study team as a direct assessment of the healthcare workers’ free-text survey responses. An additional reviewer (AH) assessed each statement for theme content and established frequency tables of theme content by demographic variables. Illustrative quotes pertaining to commonly reported themes were extracted from the data and documented.

### Patient and Public Involvement

Patients or the public were not involved in the design, or conduct, or reporting, or dissemination plans of our research.

### Ethical approval

Our study was determined to be exempt by the Icahn School of Medicine at Mount Sinai Institutional Review Board (study#20-01964).

## Results

### Study Population

Between December 23, 2020, and February 16, 2021, 2,191 healthcare workers attempted, and 2,109 (96%) completed the survey.Among participants who completed the survey, 1,933 (92%) had been offered the COVID-19 vaccine at the time of survey completion. We restricted our analysis to the group who had been offered the vaccine. Respondents were predominantly cisgender female (70%), with a median age range of 40-49 years. By profession, 23% were allied health professionals, 21% worked in administration/management, 22% in education or research, 14% were nurses, and 14% were physicians. By ethnicity, 16% of the respondents were Hispanic of any race. Among racial categories, 10% were Black, 23% Asian or Pacific Islander, and 45% White.

### Vaccination Receipt Rate

Of respondents who were offered the vaccine, 81% reported they had received a COVID-19 vaccine, 11% reported they plan to get it but had not scheduled or gone for the vaccine yet, and 8% had not received the vaccine. Table 1 presents the bivariable analysis of vaccine receipt by gender, age, race and ethnicity, HCW role, and participation in direct patient care. COVID-19 vaccine receipt was highest among males, respondents aged less than 40 years and 60 years or greater, and White respondents. By role, physicians (95%), researchers/educators (92%), and advanced practice providers (92%) were the most likely to be vaccinated, while community-based health workers were the least likely to have received the vaccine (29%). There was no difference in vaccine receipt by workers’ involvement in direct patient care. Among healthcare workers who reported receiving an influenza vaccine, 84% also received the COVID-19 vaccine compared to 44% of healthcare workers who did not receive a seasonal flu vaccine (p < 0.001).

**Table 1.**
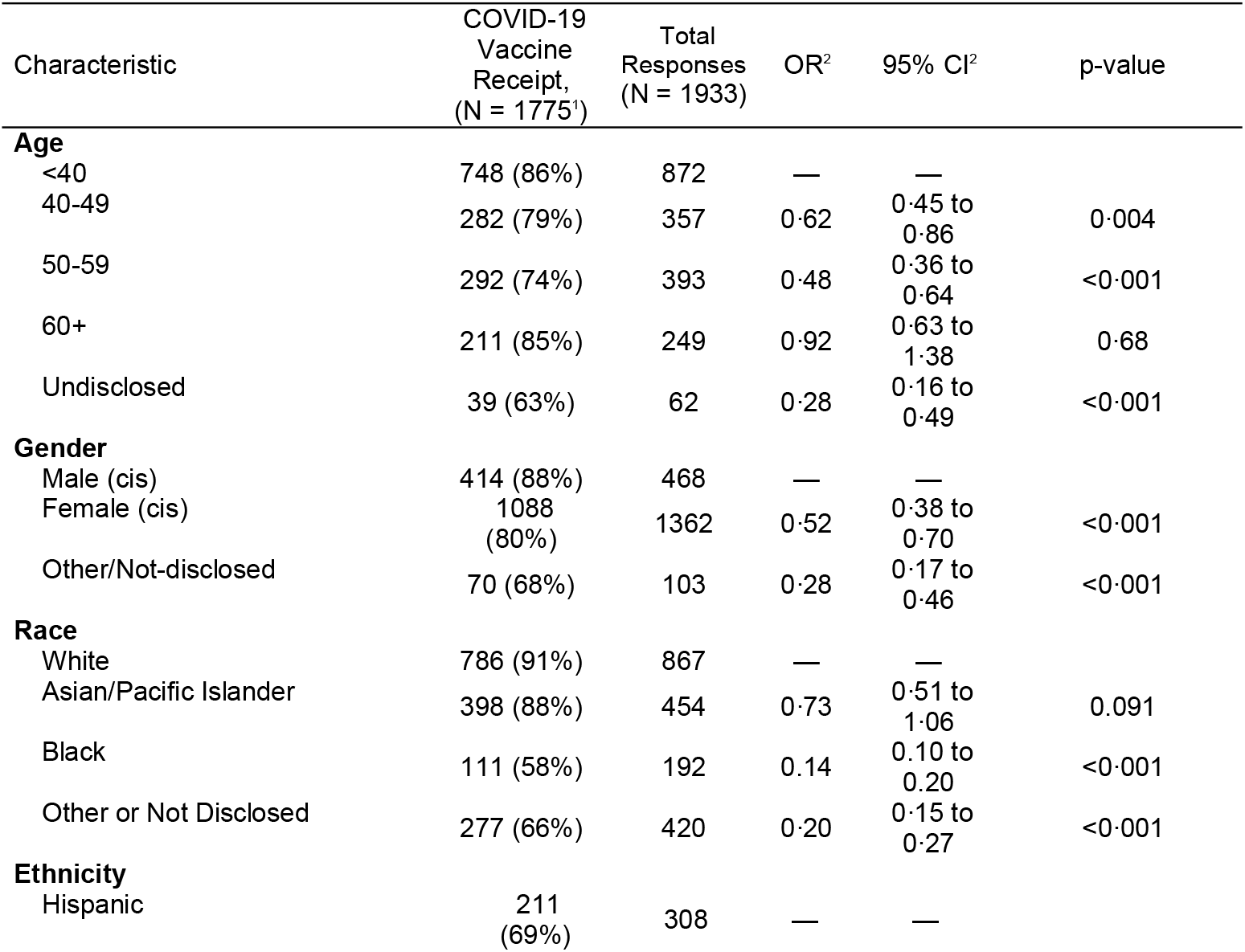
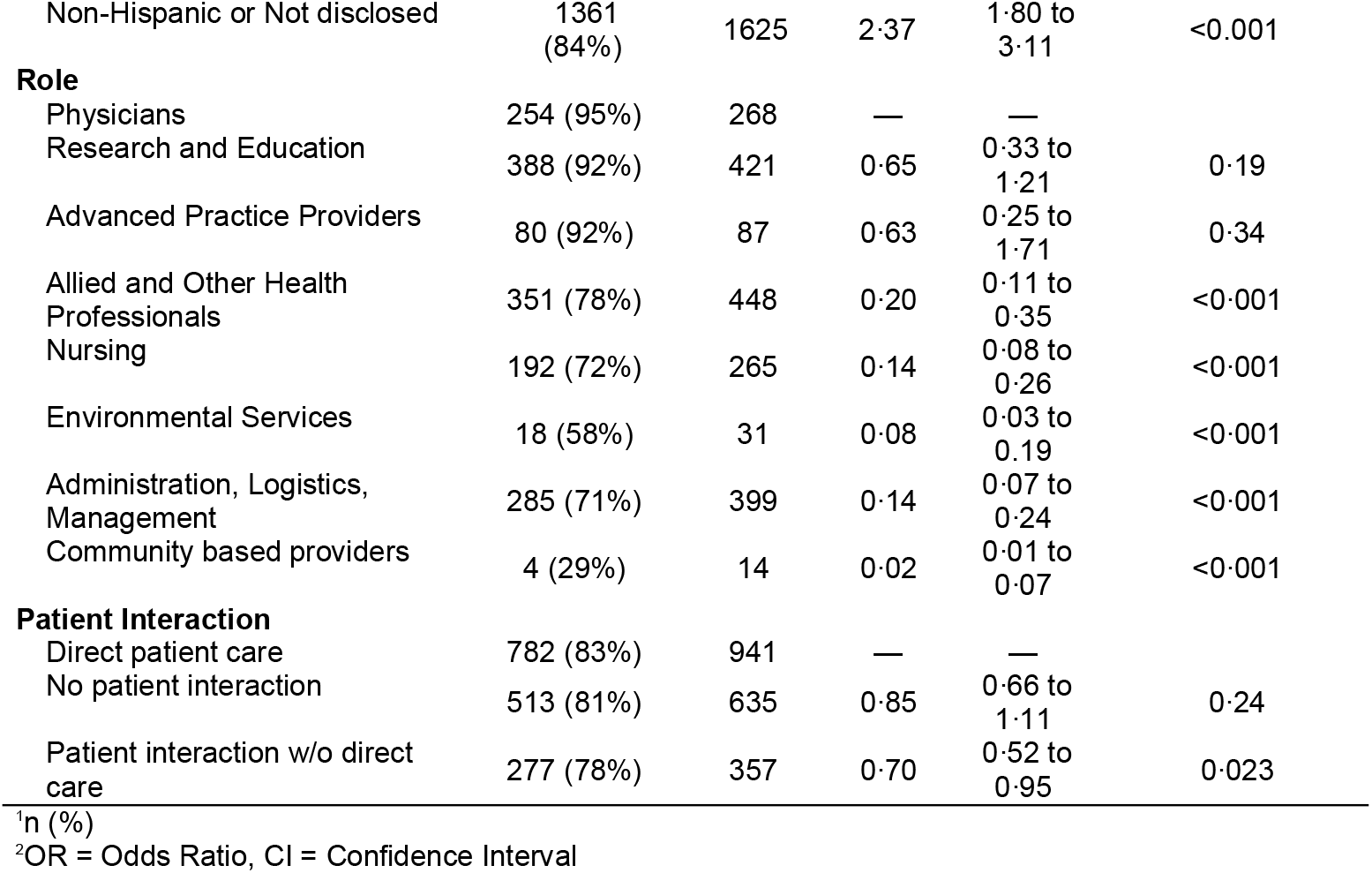
Participant Characteristics by Receipt of COVID-19 vaccine when offered.

| Characteristic | COVID-19 Vaccine Receipt, (N = 1775<sup>1</sup>) | Total Responses (N = 1933) | OR<sup>2</sup> | 95% CI<sup>2</sup> | p-value |
| --- | --- | --- | --- | --- | --- |
| **Age** |  |  |  |  |  |
| <40 | 748 (86%) | 872 | — | — |  |
| 40-49 | 282 (79%) | 357 | 0.62 | 0.45 to 0.86 | 0.004 |
| 50-59 | 292 (74%) | 393 | 0.48 | 0.36 to 0.64 | <0.001 |
| 60+ | 211 (85%) | 249 | 0.92 | 0.63 to 1.38 | 0.68 |
| Undisclosed | 39 (63%) | 62 | 0.28 | 0.16 to 0.49 | <0.001 |
| **Gender** |  |  |  |  |  |
| Male (cis) | 414 (88%) | 468 | — | — |  |
| Female (cis) | 1088 (80%) | 1362 | 0.52 | 0.38 to 0.70 | <0.001 |
| Other/Not-disclosed | 70 (68%) | 103 | 0.28 | 0.17 to 0.46 | <0.001 |
| **Race** |  |  |  |  |  |
| White | 786 (91%) | 867 | — | — |  |
| Asian/Pacific Islander | 398 (88%) | 454 | 0.73 | 0.51 to 1.06 | 0.091 |
| Black | 111 (58%) | 192 | 0.14 | 0.10 to 0.20 | <0.001 |
| Other or Not Disclosed | 277 (66%) | 420 | 0.20 | 0.15 to 0.27 | <0.001 |
| **Ethnicity** |  |  |  |  |  |
| Hispanic | 211 (69%) | 308 | — | — |  |
| Non-Hispanic or Not disclosed | 1361<br>(84%) | 1625 | 2.37 | 1.80 to<br>3.11 | <0.001 |
| **Role** |  |  |  |  |  |
| Physicians | 254 (95%) | 268 | — | — |  |
| Research and Education | 388 (92%) | 421 | 0.65 | 0.33 to<br>1.21 | 0.19 |
| Advanced Practice Providers | 80 (92%) | 87 | 0.63 | 0.25 to<br>1.71 | 0.34 |
| Allied and Other Health Professionals | 351 (78%) | 448 | 0.20 | 0.11 to<br>0.35 | <0.001 |
| Nursing | 192 (72%) | 265 | 0.14 | 0.08 to<br>0.26 | <0.001 |
| Environmental Services | 18 (58%) | 31 | 0.08 | 0.03 to<br>0.19 | <0.001 |
| Administration, Logistics, Management | 285 (71%) | 399 | 0.14 | 0.07 to<br>0.24 | <0.001 |
| Community based providers | 4 (29%) | 14 | 0.02 | 0.01 to<br>0.07 | <0.001 |
| **Patient Interaction** |  |  |  |  |  |
| Direct patient care | 782 (83%) | 941 | — | — |  |
| No patient interaction | 513 (81%) | 635 | 0.85 | 0.66 to<br>1.11 | 0.24 |
| Patient interaction w/o direct care | 277 (78%) | 357 | 0.70 | 0.52 to<br>0.95 | 0.023 |

<sup>1</sup>n (%)
<sup>2</sup>OR = Odds Ratio, CI = Confidence Interval

### Perception of COVID-19 Vaccine Benefits

Healthcare workers who agreed with statements about the COVID-19 vaccine’s importance in protecting family members were more likely to receive the vaccine compared to those who did not agree with this statement (86% compared to 25%, p <0.001). This relationship was consistent across workers with different beliefs in the vaccine’s importance in protecting community members, co-workers, and patients. Table 2 presents bivariable analysis of vaccine receipt by vaccine beliefs and risk appraisal.

**Table 2.** Risk appraisal, beliefs, and media behaviors by receipt of COVID-19 vaccine.

| Survey prompt | COVID-19 Vaccine Receipt<br>(N=1,775) | Total Responses<br>(N = 1,933) | p-value<sup>1</sup> |
| --- | --- | --- | --- |
| **Overall, I think vaccines are safe** |  |  | <0.001 |
| Agree | 1,528 (84%) | 1,810 |  |
| Neither or Disagree | 44 (36%) | 123 |  |
| **Overall, I think vaccines are effective** |  |  | <0.001 |
| Agree | 1,522 (83%) | 1,824 |  |
| Neither or Disagree | 50 (46%) | 109 |  |
| **I think vaccines are important for children to have** |  |  | <0.001 |
| Agree | 1,457 (83%) | 1,761 |  |
| Neither or Disagree | 115 (57%) | 172 |  |
| **I am worried about COVID-19 vaccine safety** |  |  | <0.001 |
| Neither or Disagree | 1,081 (92%) | 1,170 |  |
| Agree | 491 (64%) | 763 |  |
| **I am worried about possible side effects from a COVID-19 vaccine** |  |  | <0.001 |
| Neither or Disagree | 920 (92%) | 996 |  |
| Agree | 652 (70%) | 937 |  |
| **I am worried about being experimented on** |  |  | <0.001 |
| Neither or Disagree | 1,275 (90%) | 1,409 |  |
| Agree | 297 (57%) | 524 |  |
| **Getting a COVID-19 vaccine is important because it will help protect my family** |  |  | <0.001 |
| Agree | 1,535 (86%) | 1,785 |  |
| Neither or Disagree | 37 (25%) | 148 |  |
| **Getting a COVID-19 vaccine is important because it will help protect my co-workers** |  |  | <0.001 |
| Agree | 1,527 (86%) | 1,782 |  |
| Neither or Disagree | 45 (30%) | 151 |  |
| **The COVID-19 vaccine is important to prevent more deaths in my community** |  |  | <0.001 |
| Agree | 1,531 (85%) | 1,807 |  |
| Neither or Disagree | 41 (33%) | 126 |  |
| **I am at high risk for getting COVID-19 because of my job** |  |  | <0.001 |
| Agree | 1,285 (83%) | 1,549 |  |
| Neither or Disagree | 287 (75%) | 384 |  |
| **I am at high risk for complications from COVID-19 because of personal health conditions** |  |  | 0.60 |
| Neither or Disagree | 1,133 (82%) | 1,375 |  |
| Agree | 439 (79%) | 558 |  |
| **Got flu vaccine this season** |  |  | <0.001 |
| Yes | 1,508 (84%) | 1,790 |  |
| No or Don't remember | 64 (45%) | 143 |  |
| **Hours per day spent using social media in last 2 weeks** |  |  | 0.2 |
| 1+ hours | 839 (82%) | 1,018 |  |
| Under 1 hour | 733 (80%) | 915 |  |
| **Hours per day spent watching TV or digital media in last 2 weeks** |  |  | 0.3 |
| 1+ hours | 1,216 (82%) | 1,486 |  |
| Under 1 hour | 356 (80%) | 447 |  |

<sup>1</sup>Pearson's Chi-squared test

### Perception of COVID-19 Vaccine Risks

Regarding potential COVID-19 vaccine risks, 27% of respondents expressed concern about being experimented on, 40% expressed concern about vaccine safety, and 48% expressed concern about side effects. Among respondents who expressed concern about safety, 64% received the vaccine, as did 70% with concerns about side effects, compared to 92% of respondents who did not express those concerns (p < 0.001). Similarly, respondents who expressed concern about being experimented on were less likely to have received the vaccine compared to those who did not (57% compared to 90%, p < 0.001). Figure 2 presents the percent of respondents who strongly or somewhat agreed with the statement “I am worried about being experimented on”, by race and ethnicity and healthcare worker role. Among all participants, 94% of respondents agreed that “overall, vaccines are safe” and 93% received the influenza vaccine during the most recent flu season (2020-2021).

### COVID-19 Disease Risk Appraisal

Across all participants, 80% believed they were at high risk of getting COVID-19 due to their job. Healthcare workers who felt they were at high risk due to their job were more likely to have received the vaccine (83% compared to 75%, p < 0.001). Among those who disagreed that their job put them at high risk, 69% still reported vaccine receipt. Being at high risk for COVID-19 complications due to underlying health conditions did not affect vaccine uptake (79% compared to 82%, p=0.07).Healthcare workers who responded that they did not need the vaccine because they had a history of COVID-19 disease or had positive antibodies represented 6% of total participants but made up 21% of participants who did not receive the vaccine.

### Multivariable Analysis

The final multivariable model is shown in Figure 1 (See Supplementary Table 1 for LASSO coefficients). Identifying as Black was associated with a decreased vaccine receipt (odds ratio 0.38, 95% confidence interval 0.24 to 0.59). Concerns about vaccine safety and being experimented on continued to predict lower vaccine receipt (odds ratio 0.39, 95%CI 0.28-0.55 and odds ratio 0.44, 95%CI 0.31 to 0.60). Beliefs in the importance of the vaccine protecting others remained associated with higher vaccine receipt (odds ratio 2.69, 95%CI 1.93-3.74). By occupational category, a role in nursing (OR 0.37, 95%CI 0.21-0.65), administration (OR 0.46, 95%CI 0.26-0.78), or allied and other health professionals (OR 0.48, 95%CI 0.27-0.81) remained significant for decreased odds of vaccine receipt compared to physicians and advanced practice providers. Influenza vaccine receipt (odds ratio 3.57, 95%CI 2.30 to 5.56) was also associated with a higher odds of COVID-19 vaccine uptake. Higher general vaccine confidence measured by the Vaccine Confidence Index (OR 1.06, 0.99-1.14, p=0.076) was not significant in the final multivariable model.

**Figure 1.**
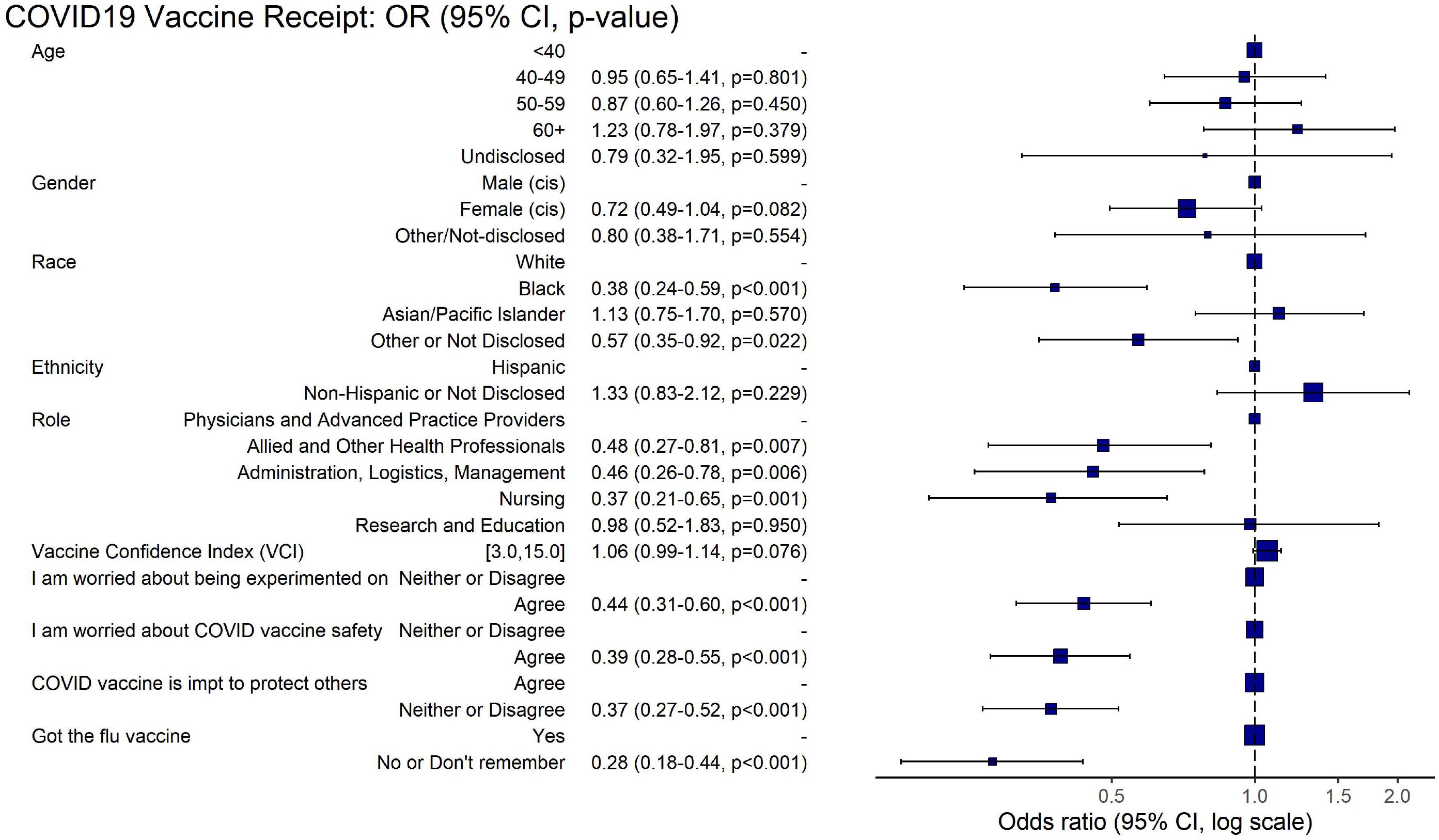
Multivariable logistic regression model on the predictors of COVID-19 vaccine receipt

**Figure 2.**
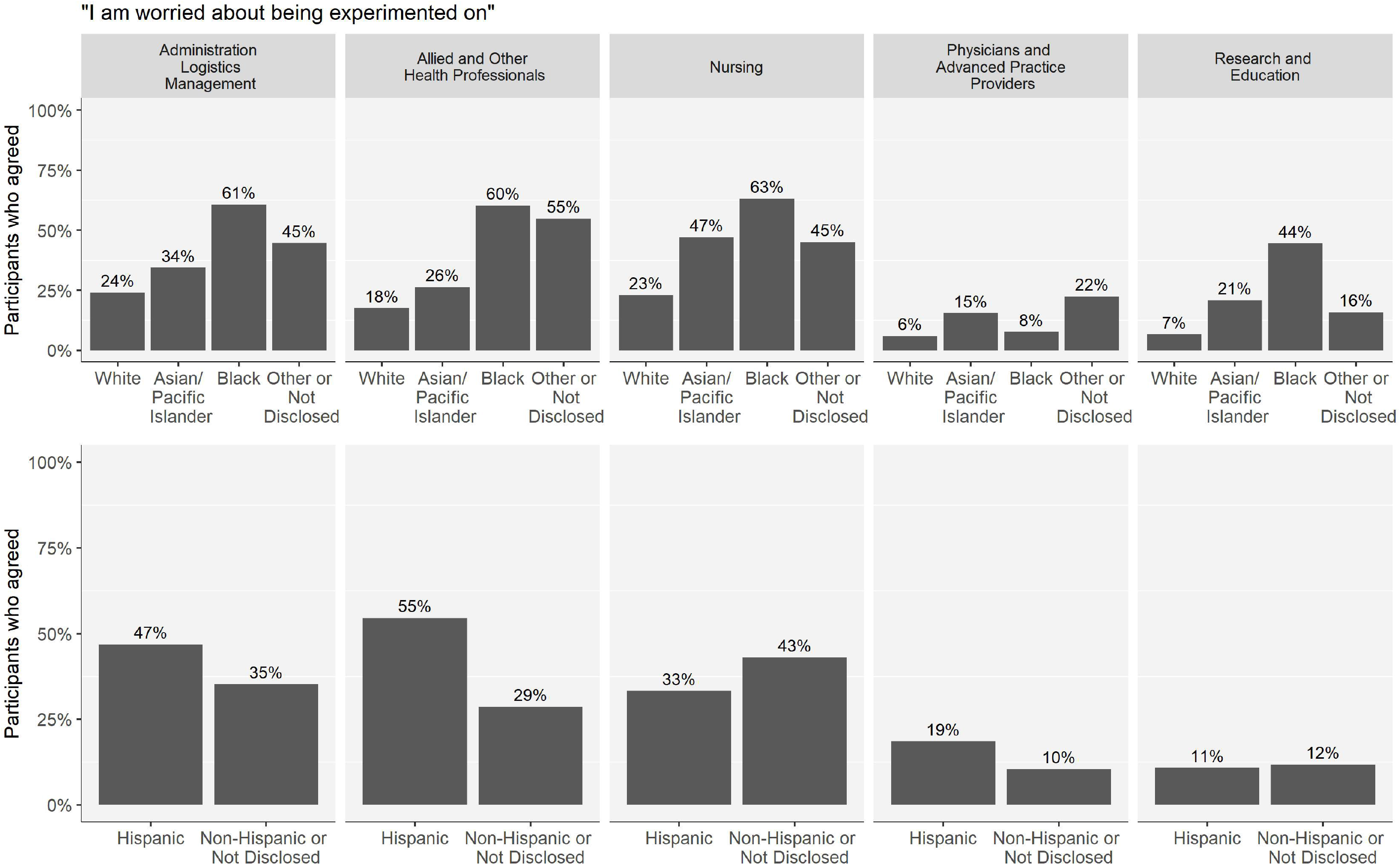
Percent of respondents strongly or somewhat agreeing with the statement “I am worried about being experimented on”, by Race and Ethnicity

### Trusted sources of COVID-19 vaccine information and social media patterns

Healthcare workers reported they were asked their opinion about COVID-19 vaccination multiple times in the past month by coworkers (73%), family members (86%), and community members or friends (73%). When asked “who do you trust MOST to give you advice on COVID-19 vaccines?”, 39% of healthcare workers chose “my primary care doctor”, followed by “federal government agencies” (28%) and “other healthcare professionals” (12%). Among healthcare workers who did not get the COVID-19 vaccine by the time of survey completion, the most trusted source was also their primary care doctor, followed by federal government agencies.

### Qualitative results

Overall, 641 respondents provided free text answers to the open-ended questions at the end of the structured survey. Of these respondents, 459 (72%) were female, 142 (22%) were male and 40 (6%) were Queer/Non-Binary, Transgender FTM, other gender, or preferred not to identify their gender. We summarize qualitative results of the free-text responses in Supplementary Table 2. The primary themes were: 1) Positive regard for the COVID-19 vaccine; 2) Mistrust in public health institutions and government; 3) Specific concerns about COVID-19 vaccines; 4) Identification of vaccine related-education needs; and 5) Suggestions for mass vaccination and distribution.

The first of the primary themes identified healthcare workers’ overall positive valence towards the COVID-19 vaccine and expressing keenly wanting to receive it. Participants’ statements of ‘positive regard’ indicated gratitude, interest in sharing positive thoughts with others, and a hope that everyone will receive the vaccine as soon as possible. The theme of general mistrust in public health institutions and government around the COVID-19 pandemic overall, and in various considerations related to the vaccine, also emerged. The theme of mistrust was identified in only 8% of statements. However, statements aligning with the theme of mistrust in the vaccine were more frequently cited among healthcare workers who identified as Black, and Black healthcare workers provided reasons for hesitancy more frequently than other racial/ethnic groups. The proportion of themes expressed by healthcare workers by race and ethnicity are shown in Supplementary Table 3. Reasons for hesitancy included uncertainty about long term side effects, needing more time to “wait and see” what happens when others take the vaccine. Participants highlighted the historical medical experimentation on minority groups, political involvement in the COVID-19 pandemic response overall and in the development and distribution of the vaccine. For the theme ‘Specific concerns about COVID-19 vaccines’ participants described a range of reasons for their concerns in obtaining the vaccine. Common specific concerns expressed included side effects, uncertainty around the need for vaccine if previously infected, reproductive health considerations, and a concern that the vaccine was developed too quickly. This theme was also represented by desires to be vaccinated after more data and experience with the vaccine became available. Participants ‘suggestions for mass vaccination and distribution’ included logistical improvements for vaccine sites, transparency in distribution, providing incentives for vaccination, and ideas regarding the order of eligibility.

In response to the prompt for suggested tools and resources participants had a variety of suggestions for what is needed regarding vaccine education and confidence promotion. The identification of educational needs was the most frequently endorsed theme. Fifty percent of healthcare workers who provided a free text response identified a need for more information pertaining to the COVID-19 vaccine. Participants suggested efforts related to methods of communication and identified trusted sources for dissemination of information. Statements reflected a need for infographic materials to explain how the vaccines work. Healthcare workers indicated a need to explain how the vaccines work, in particular the mRNA vaccines. Healthcare workers stated their continued interest in receiving updates of COVID-19 vaccination information via easy to access academic journals and webinars. Many healthcare workers expressed an interest in greater use of health information technology to disseminate COVID-19 vaccination information about side effects, efficacy, and distribution.

## Discussion

We found that while COVID-19 vaccine uptake among healthcare workers in our study is the norm—81% of respondents received the vaccine—there were important inequities in vaccine receipt by race, gender, age, and healthcare worker role. Black, Hispanic and female participants were less likely to have received the vaccine, while physicians, advanced practice providers, and researchers/educators were more likely to have received the vaccine. Our study, occurring in the immediate two-month period following vaccine rollout among healthcare workers, is one of the first studies to report COVID-19 vaccine receipt rather than intent, and the first to investigate the association of COVID-19 vaccine receipt with vaccine confidence and perceptions of vaccine risks and benefits among healthcare workers.

Our results on race support findings from earlier research that examined COVID-19 vaccine intent prior to vaccine availability^22-28^. Similarly, many published studies have found men were more willing to accept a potential COVID-19 vaccine than women^1,23,25^ though one found the opposite.^29^ Consistent with early reports of COVID-19 vaccination rates in the United States, we found Black and Hispanic participants were less likely to receive the COVID-19 vaccine at the time of our survey. The Centers for Disease Control and Prevention reported that among persons who received the vaccine, the proportion who identified as Black was lower than would be expected based on eligibility.^30^ Similarly, in New York City at the time of our study, Black New Yorkers made up 12% of vaccine recipients, but 24% of the population.^31^ Both reports were limited by incomplete data collection for race and ethnicity, but in our sample 94% of respondents reported race/ethnicity data.

We found higher perceived benefits of COVID-19 vaccination for protecting others had the largest effect size in predicting vaccine receipt. Lower perceived vaccine harms including safety and being experimented on also predicted vaccine receipt. Higher perceived benefits of COVID-19 vaccination, higher perceived risk of COVID-19 disease, and more positive attitudes towards vaccination have been associated with greater intent to vaccinate in the general US population.^1,23,25^ We found no difference in vaccine uptake based on whether respondents had patient-facing roles. This may be explained by healthcare workers’ confidence in the protection provided by personal protective equipment (e.g., masks, face shields) or that healthcare workers have high perceived COVID-19 risk regardless of direct patient interaction. We do not have more refined data, however, as to whether site of patient care – e.g., in home or community settings versus in clinic or hospital facilities – might moderate perceived risk.

A prior study investigating COVID-19 beliefs, vaccine intent, and race found that beliefs mediate the association of race and vaccine intent.^32^ While Black respondents in their study had lower vaccine intent and lower pro-vaccine beliefs, race was not significant when controlling for pro-vaccine beliefs. Here, Black healthcare workers had lower COVID-19 vaccination rates even after accounting for safety concerns and pro-vaccine beliefs in the multivariable model. Vaccine intent does not always translate to vaccine receipt^33^ and this effect may be greater among Black healthcare workers. The effects of systemic racism, the history of research abuses among people of color in the United States, and the lived experience of mistreatment in healthcare systems likely all contribute to lower vaccination rates.^22,34,35^ Increasing access to evidence-based information and implementing various forms of reflective listening with marginalized communities who are less likely to take the vaccine may decrease mistrust among healthcare workers critical to achieving a more equitable vaccine response. Additionally, while all healthcare workers in this study had been offered the vaccine in their workplaces, there may have been racial differences in actual access (e.g., ability to take time off duty to wait for the vaccine) that our study was not designed to detect.

A strong physician recommendation in favor of vaccination has been shown to positively influence vaccine decision making for many immunizations, and COVID-19 appears no different.^36,37^ In our study, healthcare workers reported their primary care doctor was the most trusted source for advice on COVID-19 vaccination and this held true across race and vaccine receipt. Our findings also point to the influence that all healthcare workers can have in discussing COVID-19 vaccination within social networks of co-workers, family and community members. Further research is needed in developing strategies for healthcare workers to listen to, respond, and address issues related to vaccine equity and uptake in the context of systemic racism and barriers to care.

### Limitations

The high rate of COVID-19 vaccine acceptance in our population may be the result of response bias, where healthcare workers with favorable vaccine attitudes were more likely to complete the survey. Our study may have some limits in generalizability given the setting in New York City and a substantial representation from an academic health system. Additionally, Black and Hispanic respondents were underrepresented in our survey, comprising 10% and 16% of our survey population respectively, whereas approximately 20% of the healthcare workers in our systems’ hospitals identify as Black and 18% identify as Hispanic.

We chose explicitly to use *receipt* as the primary outcome, rather than *intent*. As such, participants who indicated they “planned” to receive the vaccine were categorized for the primary outcome along with those who stated that they did not plan to. We did review the differences between these two groups and assessed differences over time (see Supplementary Figure 5). Those who stated an intent may have had perceptual or belief reasons for delaying, as several participants expressed in their free text responses. Additionally, while the COVID-19 vaccines were made *available* to employees at the time of the survey, many may have had *access* issues, which our survey was not designed to assess. The 11% of respondents in our survey who reported they planned to get the vaccine but had not scheduled or gone for their vaccine yet may have experienced access issues. We recognize that access plays an important role in vaccine uptake and future studies should investigate inequities in access to vaccines and to accurate information. Intervention design targeting marginalized groups of healthcare workers and others will need to account for the dynamic interplay between vaccine access and ease of access, respectful service delivery, and vaccine behaviors.

## Conclusion

In this study of 1,933 healthcare workers during the initial COVID-19 vaccine rollout in New York City, our data demonstrated high overall receipt and confidence. Beliefs in the COVID-19 vaccine’s importance in protecting others were the strongest independent predictors of vaccine receipt. Even among healthcare workers with concerns about safety, side effects or being experimented on, over 50% did receive the first dose of vaccine, suggesting a potential pathway for intervention among these workers. Beyond beliefs specific to COVID-19 vaccination, overall vaccine confidence on a standardized instrument was high.

Our study also demonstrated striking inequities in COVID-19 vaccine receipt. Black healthcare workers, adjusting for occupation and other factors, were less likely to receive the COVID-19 vaccine. A quarter of healthcare workers expressed concerns about being experimented on, particularly among marginalized groups. Addressing mistrust in public health and healthcare related to systemic racism will be critical in achieving a more equitable vaccine response.

## Data Availability

Data available by request to authors.

## Supplementary Tables and Figures

Supplementary Figure 1. Percent of respondents who were asked their opinion about COVID-19 vaccination by coworkers, family and community members

Supplementary Figure 2. Vaccine Confidence Index by Race and Ethnicity

Supplementary Figure 3. Multivariable analysis including only demographic variables

Supplementary Figure 4. Additional COVID-19 vaccine specific concerns by race (top) and Hispanic ethnicity (bottom).

Supplementary Figure 5. Vaccine receipt over time of the survey.

Supplementary Table 1. Lasso Regression of participant characteristics and beliefs

SupplementaryTable 2. Qualitative Analysis Summary Table of Free Text Survey Responses

Supplementary Table 3. Themes expressed in healthcare workers’ free text responses about COVID-19 vaccination by race and ethnicity

## Acknowledgments

We would like to acknowledge all the anonymous healthcare employees who took part in our survey, and thank them for their care, strength and commitment during this difficult pandemic. We would also like to thank the COVID-19 Research Unit at Elmhurst Hospital (CURE-19), along with its sponsors, the Global Health Institute at NYC Health and Hospitals / Elmhurst Hospital Center and the Arnhold Institute for Global Health at the Icahn School of Medicine at Mount Sinai.

## Contributorship

ABo, AF, AG, AH, CH, JJ, JK, SL, DM, SM, JM, KO, SP, DP, AFa, ARo, LS, JL, RV conceptualized the study, ABa, ABo, ND, CG, CH, JK, SL, DM,KO, NS, LS developed the methodology, ABo, CH, JJ, DM, SM, KO, SP, ARa, NS, LS developed the survey design, AH, JK, SL, DM,SP, NS, LS, NV,NI conducted dissemination, DD, CH,SP, ARa, LS, JL, RV curated the data, IN, SP, NV conducted data validation, AC,JK,RM, DM, JM, SP, ARa, NS, LS analyzed the data, DM, ARa created data visualizations, AC, AG, DM, KO, ARa, LS were responsible for project administration, KO, DM, AF, CH, LS, AH, NI wrote the original draft. All authors reviewed and edited the manuscript, provided relevant intellectual input, and read and approved the final manuscript. KO and DM have primary responsibility for the final content. KO will act as guarantor. The corresponding author attests that all listed authors meet authorship criteria.

## Ethics approval

This study received exemption determination from the Institutional Review Board at the Icahn School of Medicine at Mount Sinai (#20-01964).

## Data sharing

Data available by request to authors.

## Dissemination declaration

We plan to disseminate the results of our study to healthcare workers and stakeholders at participating institutions using infographics, summary reports and presentations.

## Copyright

The Corresponding Author has the right to grant on behalf of all authors and does grant on behalf of all authors, an exclusive licence (or non exclusive for government employees) on a worldwide basis to the BMJ Publishing Group Ltd to permit this article (if accepted) to be published in BMJ editions and any other BMJPGL products and sublicences such use and exploit all subsidiary rights, as set out in our licence.

## Transparency statement

The lead author, affirms that the manuscript is an honest, accurate, and transparent account of the study being reported; that no important aspects of the study have been omitted; and that any discrepancies from the study as planned (and, if relevant, registered) have been explained.

