## Supplemental Table 1 for "Factors associated with COVID-19 vaccine receipt at two integrated healthcare systems in New York City: A Cross sectional study of healthcare workers"

| **Predictor** | **Coefficient** |
| --- | --- |
| (Intercept) | 0.40459964 |
| (Intercept) | . |
| Age 40-49 | . |
| Age 50-59 | . |
| Age 60+ | . |
| Age Undisclosed | . |
| Race – Black | -0.48840025 |
| Race – Asian/Pacific Islander | . |
| Race – Other or Not disclosed | -0.34706673 |
| Ethnicity – Hispanic | . |
| Gender - Female (cis) | . |
| Gender - Other/Not-disclosed | . |
| VCI | 0.04761949 |
| Role – Allied and Other Health Professionals | . |
| Role – Administration, Logistics, Management | . |
| Role – Nursing | . |
| Role – Research and Education | . |
| Worried about safety – Agree | -0.74808485 |
| Worried about side effects – Agree | . |
| Worried about being experimented on – Agree | -0.78536879 |
| At high risk of COVID due to job – Neither or Disagree | -0.11697236 |
| Vaccine important to protect others – Agree | 0.60529066 |
| Got the flu vaccine - Yes | 0.90035387 |

**Supplementary Table 1. Lasso Regression of participant characteristics and beliefs**
