## Supplemental Table 2 for "Factors associated with COVID-19 vaccine receipt at two integrated healthcare systems in New York City: A Cross sectional study of healthcare workers"

| **Primary Theme** | **Description** | **Exemplar Quote** |
| --- | --- | --- |
| Positive regard for the COVID-19 vaccine | Many participants shared positive regard on the COVID-19 vaccine and expressed wanting to receive it. | *‘The benefits of the vaccine outweigh the risks of catching this deadly disease.’* |
| Mistrust in public health institutions and government | Participants expressed a general mistrust around the COVID-19 pandemic overall and in various considerations related to the vaccine. | *‘I don't trust it. I don't trust the government. They have always tried to hurt and use African Americans as test subjects.’* |
| Specific concerns about COVID vaccines | Participants describe a range of reasons for their hesitancy in obtaining the vaccine. | *‘I need to wait and see if there are any long/short term side effects from taking a vaccine for Covid.’* |
| Identification of vaccine related-education needs | Participants had suggestions for what is needed regarding vaccine education and confidence promotion. | *‘I think the information provided about vaccination should be in as clear language as possible so as not to confuse those without medical/science backgrounds. As more information about immunity conferred by the vaccine becomes available it should also be provided to help people understand the vaccination process.’* |
| Suggestions for mass vaccination and distribution | Participants provided suggestions related to mass distribution. | *‘The process for receiving a vaccine was unorganized and it was confusing to know who was eligible or not. better guidelines should have been in place to prevent misinformation and ensuring those who were supposed to be vaccinated go [get] vaccinated.’* |

**Supplementary Table 2. Qualitative Analysis Summary Table of Free Text Survey Responses**
