## Supplemental Table 3 for "Factors associated with COVID-19 vaccine receipt at two integrated healthcare systems in New York City: A Cross sectional study of healthcare workers"

|  | White  (n=256) | Black  (n=79) | Asian/  Pacific Islander  (n=170) | Other Race/Not Disclosed  (n=136) | Hispanic  (n=93) | Overall  (n = 641) |
| --- | --- | --- | --- | --- | --- | --- |
| Positive Regard | 60 (23%) | 16 (20%) | 33 (19%) | 21 (15%) | 14 (15%) | 130 (20% |
| Mistrust | 19 (7%) | 12 (15%) | 9 (5%) | 13 (10%) | 7 (8%) | 53 (8%) |
| Reasons for Hesitancy | 19 (7%) | 9 (11%) | 8 (5%) | 8 (6%) | 4 (4%) | 44 (7%) |
| Education | 168 (66%) | 33 (42%) | 70 (41%) | 52 (38%) | 44 (47%) | 323 (50%) |
| Suggestions for Mass Vaccination/Distribution | 51 (20%) | 25 (32%) | 36 (21%) | 17 (13%) | 18 (19%) | 129 (20%) |
| Specific Concerns | 68 (27%) | 21 (27%) | 23 (14%) | 29 (21%) | 19 (20%) | 141 (22%) |

**Supplementary Table 3. Themes expressed in healthcare workers' free text responses about COVID-19 vaccination by race and ethnicity**
