## Supplementary figures and images for "Factors associated with COVID-19 vaccine receipt at two integrated healthcare systems in New York City: A Cross sectional study of healthcare workers"

### Supplemental Figure 1

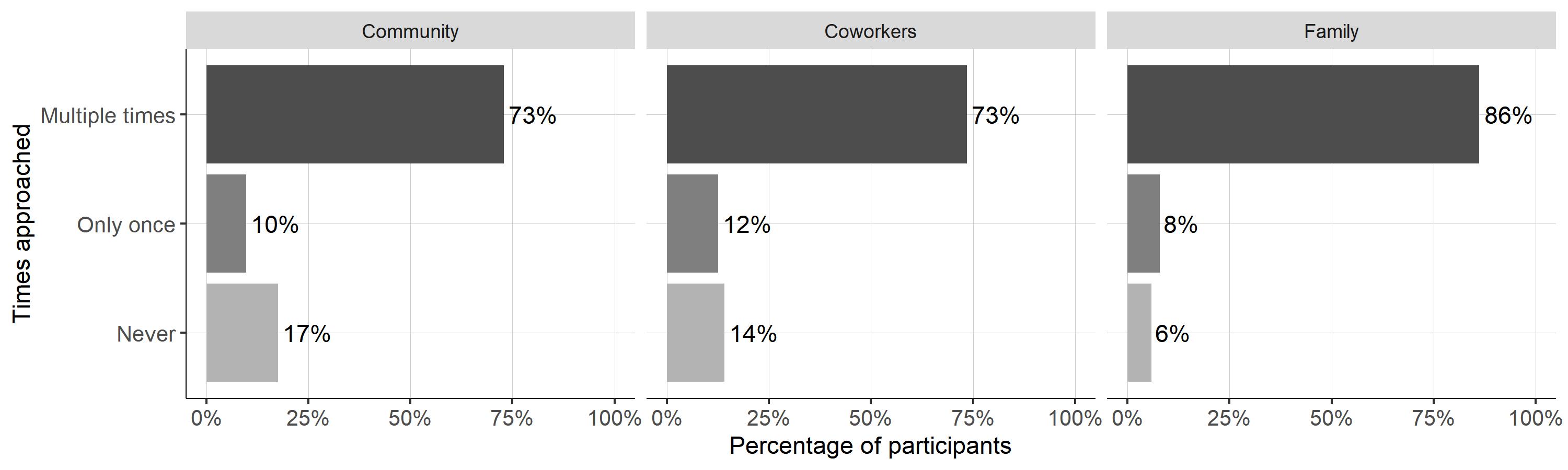

### Supplemental Figure 2

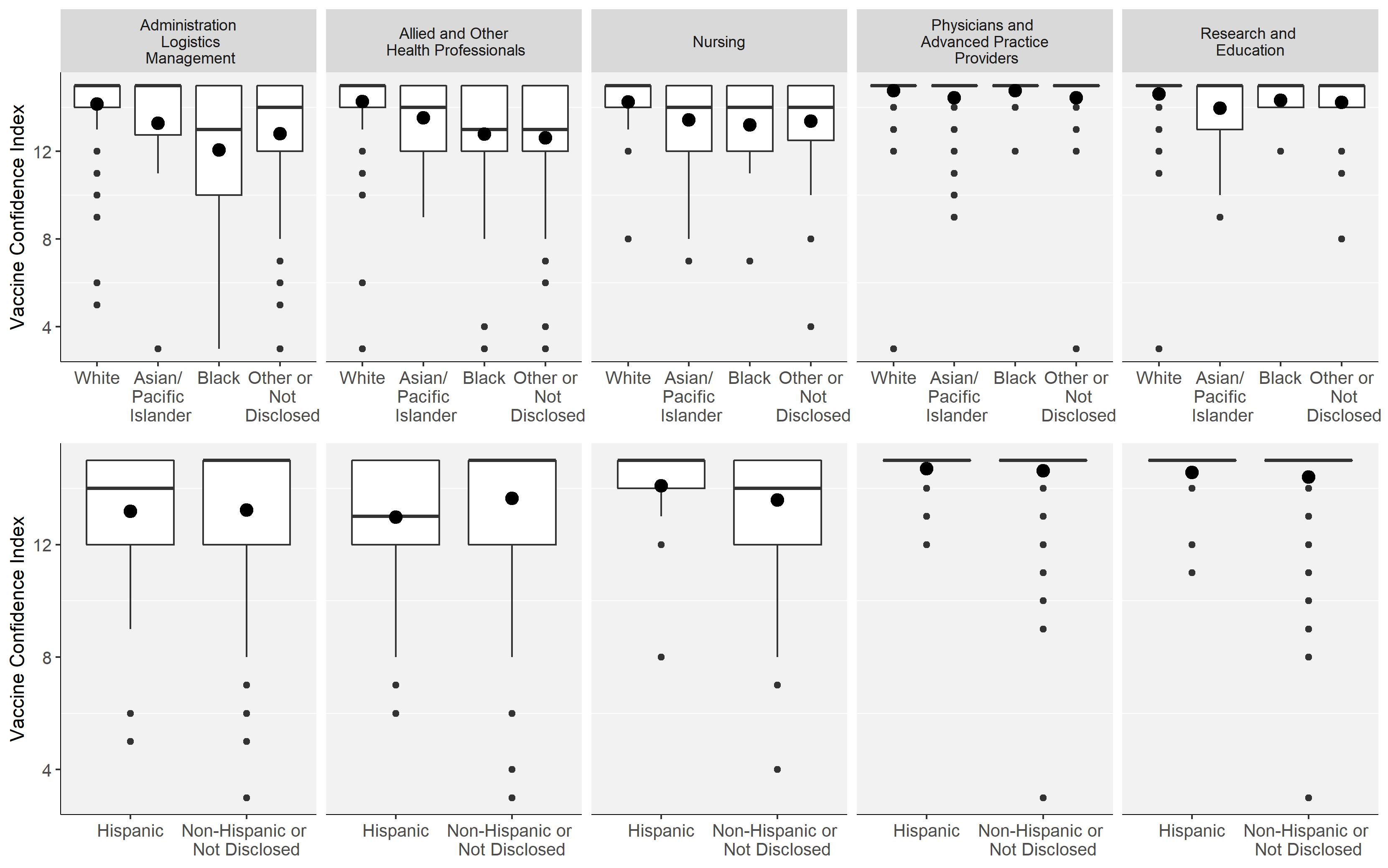

### Supplemental Figure 3

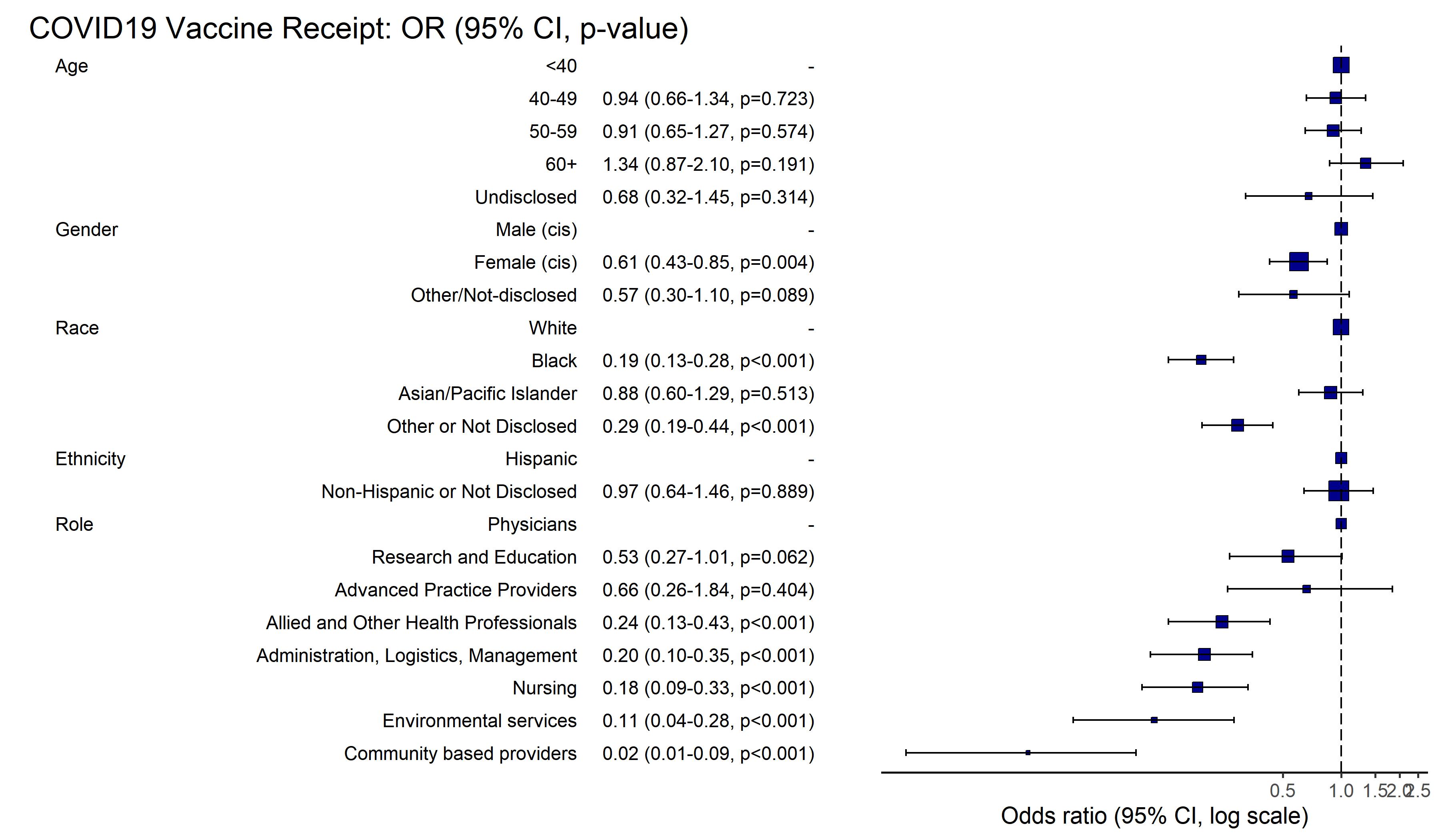

### Supplemental Figure 4

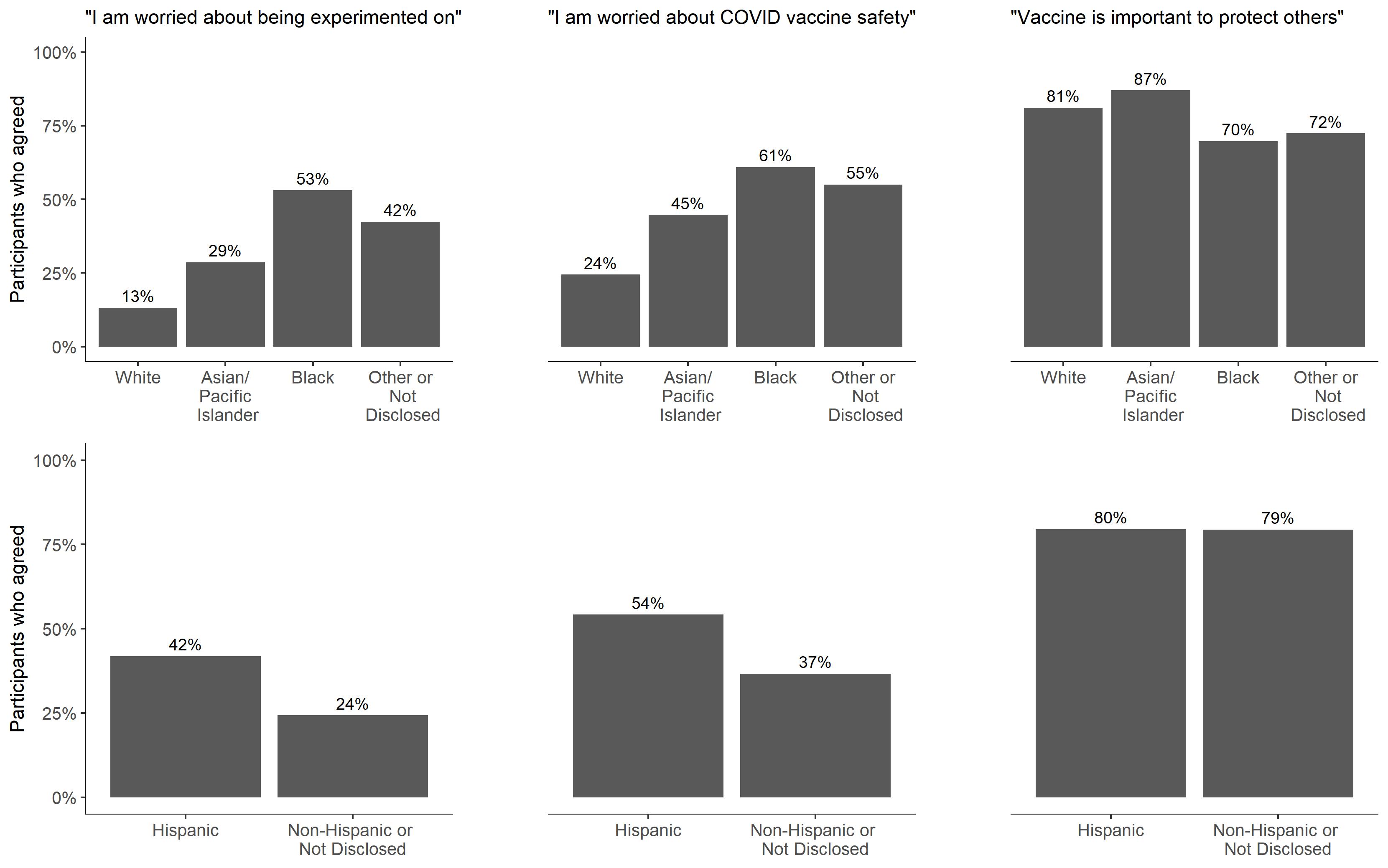

### Supplemental Figure 5

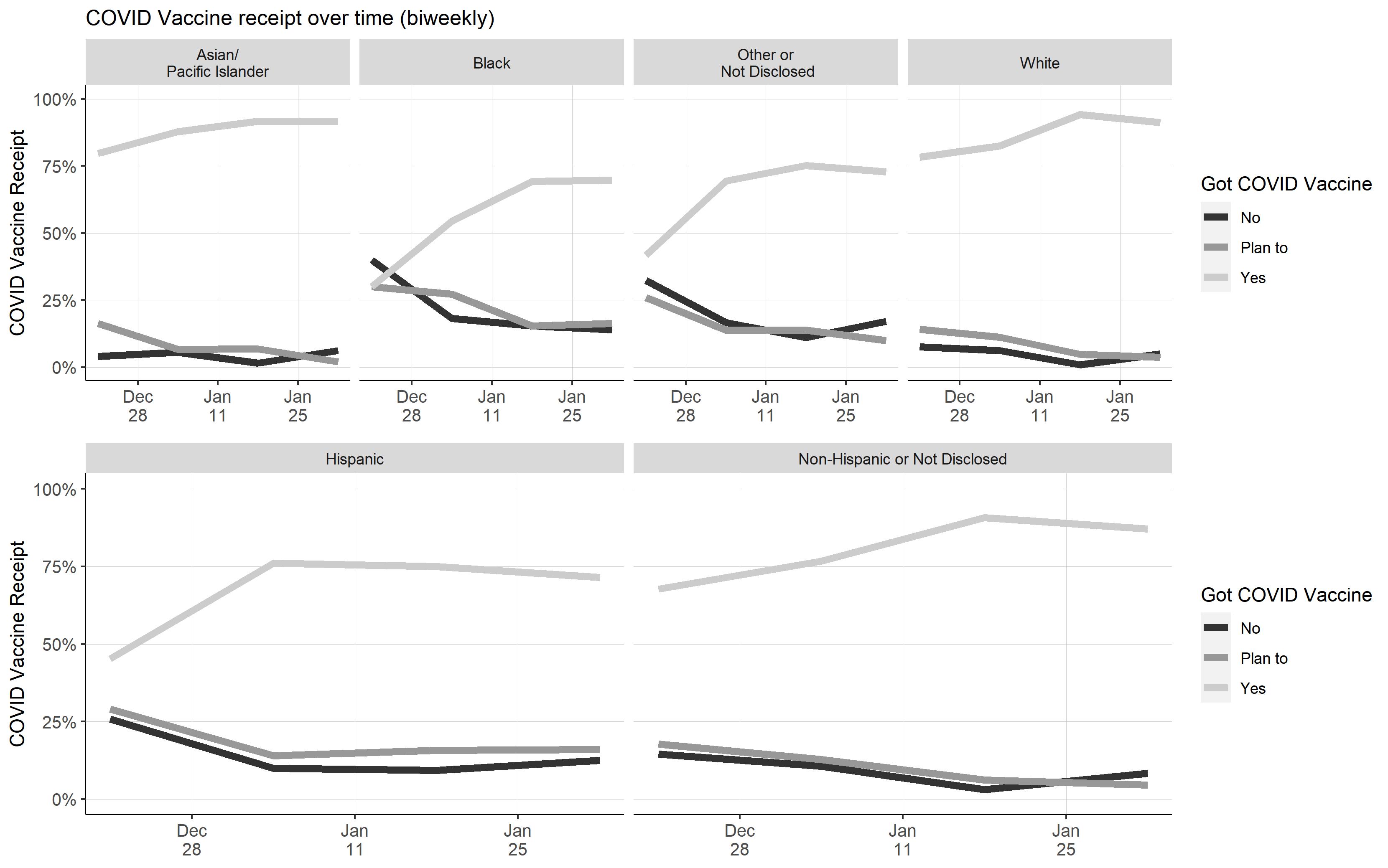
